# Assessing Male Partners’ Attitudes, Perceived Subjective Norms and Perceived Behavior Control Towards Their Involvement in Antenatal Care Clinic with Their Pregnant Women in Dodoma Urban Municipal

**DOI:** 10.1101/2024.04.03.24304992

**Authors:** Kaiza Benson Mkahala, Tienyi Mnyoro Daniel

**Affiliations:** The University of Dodoma

**Keywords:** Attitudes, perceived subjective norms, Perceived behavior control, Male involvement

## Abstract

**Background:** According to WHO, 2018 Males partners needs to know and be aware of number of antenatal visits that her pregnant woman should have, also they must know investigations and screenings which a pregnant women must undergo such as Urinalysis, HIV/AIDS screening to both male and female. Male partners’ attitudes, subjective norms, and perceived behavior control toward their involvement in ANC clinics with female partners can influence male involvement.

**Objective:** The main goal of this study was to assess male partners’ attitudes, perceived subjective norms and perceived behavior control towards their involvement in ANC clinic with their pregnant women at Dodoma urban municipal.

**Methodology:** This was a quantitative hospital-based analytical cross-sectional study, with a sample size of 377 conducted at Makole Health Centre in Dodoma for a maximum period of two weeks on June 2022. Simple random sampling was employed in choosing a sample to represent the study population. Data was collected using both self and interviewer-administered questionnaires. On data analysis was by using the Statistical Package for the Social Sciences (SPSS) version 20 to generate descriptive statistical information presented in the form of tables.

**Results:** In attitude, (183,89.7%) of male partners have positive attitude, while the remaining (21,10.3%) have negative attitude, on perceived subjective norms, (160,78.4%) of male partners have positive perceived subjective norms, while the remaining (44,21.6%) have negative perceived subjective norms, and on perceived behavior control, (163,79.9%) of male partners have positive perceived behavior control, while the remaining (41,20.1%) have negative perceived behavior control towards their involvement in ANC visits with their pregnant women. After adjusting the confounders, only perceived behavior control had factors influencing positive perceived behavior control towards men involvement in ANC which were: - distance from nearby health facility [Less than 1km (AOR=0.440 at 95% CI = 0.201-0.965)] and religion [Christian (AOR=0.459 at 95% CI=0.220-0.957)].

**Conclusion:** From this study, Male partners’ attitudes, perceived subjective norms and perceived behavior control towards their involvement in ANC visits were influenced by level of education those with tertiary level (colleges and universities) have positive perceived subjective norms compared to others.

## INTRODUCTION

### Background

According to the 2016 WHO ANC model, a minimum of eight ANC visits should be arranged, with the first contact occurring in the first trimester (up to 12 weeks of pregnancy), two contacts in the second trimester (at 20 and 26 weeks of pregnancy), and five contacts in the third trimester (at 30, 34, 36, 38 and 40 weeks) (Messages et al., 2018). The word "contact" has been used instead of "visit" in this model because it implies an active link between a pregnant woman and a health-care practitioner, which the word "visit" does not imply (Messages et al., 2018).

At least one ANC appointment with a qualified health professional is attended by 85 percent of pregnant women worldwide, and 58 percent attend at least four ANC visits (Kabanga et al., 2019). However, ANC use differs between and among countries: Despite the fact that male involvement improves obstetric care seeking behavior, male involvement remains unacceptably low in poor nations (Gibore et al., 2019). Male partners’ attitudes, subjective norms, and perceived behavior control toward their involvement in ANC clinics with female partners can influence male involvement in maternity services consumption (Moshi et al., 2020).

This study aimed on assessing male partners’ attitudes, perceived subjective norms, and perceived behavior control towards their involvement in ANC visits with their pregnant women Even if a lot of research have been done on the men partners involvement in antenatal care with their pregnant women, but in reference to clinical practice done by Bachelor of science in nursing 3(BScN3) from February to October 2021, at the Sokoine Health Centre there were few number of men adhering in the antenatal care clinic with their pregnant women, and no any research paper has published on the male partners’ attitudes, perceived subjective norms and perceived behavior control towards their involvement in antenatal care clinic with their pregnant women at Singida.

## MATERIALS AND METHODS

### Study setting, period and design

This was hospital facility-based analytical cross-sectional study conducted at Dodoma urban municipal at Makole Health Centre for a maximum period of two weeks on June 2022.

### Study Population

The study participants included all walking-in male partners participating in antenatal care visits with their female partners at a time of data collection. All male partners who declined to be among the participants of the study were excluded.

### Participants recruitment

Dodoma President’s Office, Regional Administration and Local Government Tanzania (PO-RALG) provided a letter of authorization. Participants in the study provided both verbal and written agreement after being informed of the goals and methods of the study and guranteed of their right to withdraw from it at any time. The findings were from analytical crossectional study conducted at Dodoma Urban District on which the data were obtained from 204 members of the community at Makole Health Centre who after being elaborated about the objective of the study voluntarily consented to be a participant of the study and hence to be interviewed satisfied the inclusion criteria and by completely filed the given questionnairre.

### Flow of recruitment including response rate

After thorough explaining the objectives of a study to both individual, society and national level, male partners responded at a rate of 32 per day after one week of data collection, with 204 participant recruited and no non-response rate noted.

**Figure 1:**
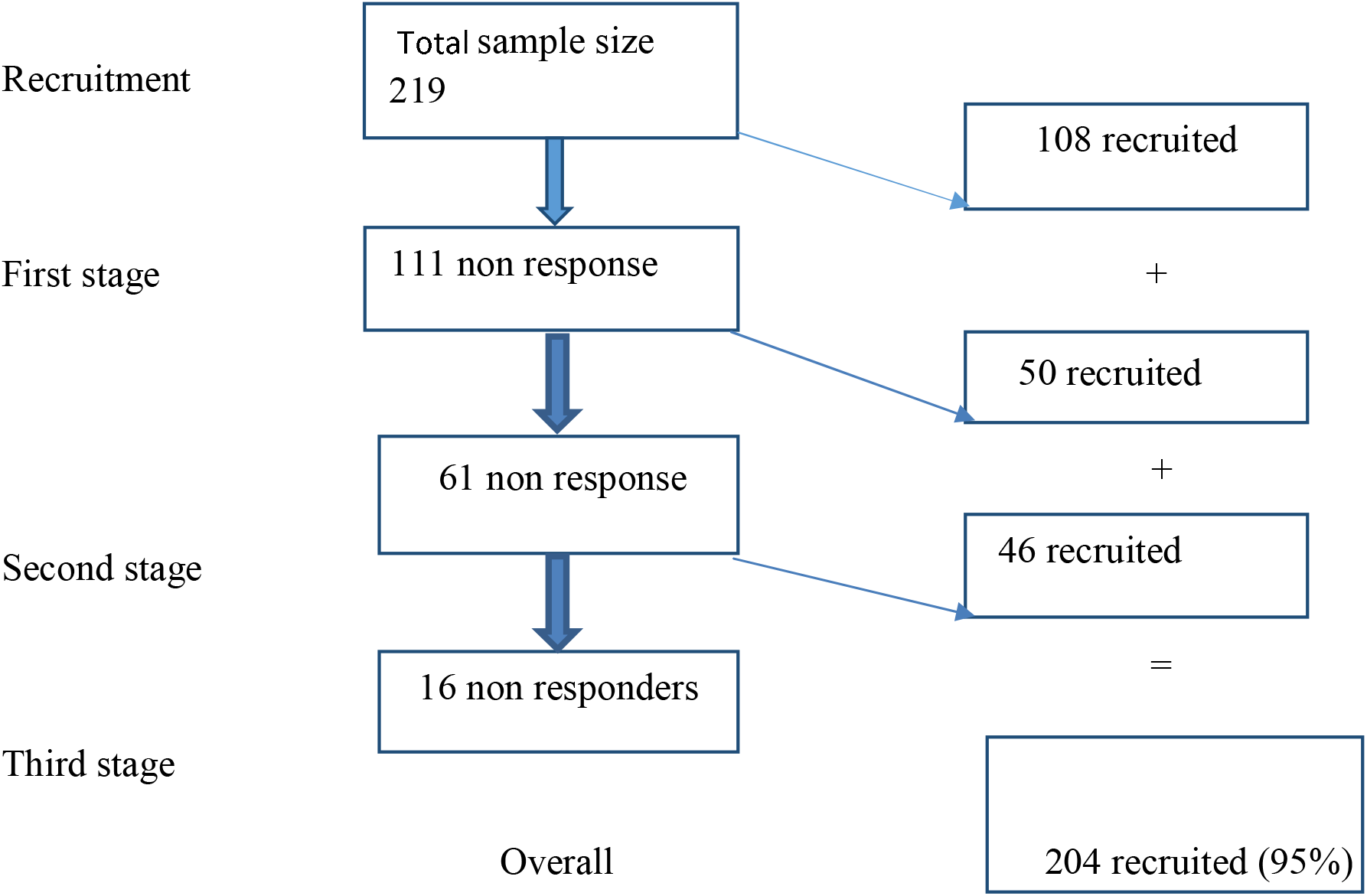
Flow chart of the recruitment and participants

### Sample size calculation and sampling technique

#### Sample size determination

The following formula was used in estimating sample size:

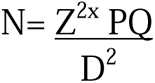

Where;

N= Desired sample size (when the population > 10000)

Z= Standard normal deviate; usually set at 1.96 which correspond to 95% confidence level

P= Proportion in a target population estimated to have a particular characteristic, 56.9% (Gibore et al., 2019), standard value of 0.569

Q= Proportion in a target population not having a particular characteristic (Q=1-P), where 1-0.569= 0.431

D= Degree of accuracy required, usually set at 0.05 level (5%) N= 377

The estimated sample size is 377

Due to limited time and resources, the estimated sample size used was 204

### Sampling technique

Simple random sampling was employed in selecting a sample to represent the study population, where every participant/respondent had an equal chance to participate in the study. Male partners were selected randomly at antenatal visits in Makole Health Centre in Dodoma.

### Data Collection and Management

The study employed both interviewer and self-administered questionnaire during conduction at Makole Health Centre, Dodoma Urban Municipal. The participants who were selected in this study were given detailed information related to the study and then asked for their consent. After clarification on each and every detail about the needs of the study, respect and confidentiality each participant who consented to be involved in the study was given a questionnaire for self-reading the questions and then providing the responses and those who were incapable of answering the questionnaire by themselves were asked the questions orally by the researcher and responses were filled on the questionnaire respectively. Participants were given at least 10-15 minutes to complete the questionnaire before collecting them back.

The questionnaire was adopted from the study of exploring factors influencing pregnant Women’s attitudes, perceived subjective norms and perceived behavior control towards male involvement in maternal services utilization (Moshi et al,2020).

According to Moshi et al., (2020) the theory of planned behavior questionnaire guide was used to guide the development of the questionnaire. The questionnaire was translated into Swahili and was pretested before actual administration.

One research assistant was recruited, trained, and participated in data collection.

A tool (questionnaire) had two parts; **the social demographic characteristics** and **a Likert scale** where respondents will be supposed to *strongly agree, agree, neutral, disagree, and strongly disagree.* The Likert scale is subdivided into three subparts of the statements in the Likert scale which are; i) male partners’ attitudes towards their involvement questions ii) male partners’ perceived subjective norms towards their involvement questions iii) male partners’ perceived behavior control towards their involvement in ANC visits.

### Attitudes of male partners towards their involvement has five Likert scale statements, which are

➢ If male partner participates in setting a part of the skilled birth attendant, he is doing a good and beneficial thing,
➢ If a male partner accompanies his pregnant female partner during antenatal clinics, he is doing a good and beneficial thing,
➢ If male partner tests for HIV with his female partner during pregnancy, he is doing a good and beneficial thing,
➢ If male partner accompanies his female partner during childbirth, he is doing a good thing which is beneficial, and
➢ If male partner accompanies his female partner for postnatal checkups, he is doing a good and beneficial thing.

### Likert scale statements involved in measuring perceived subjective norms towards male involvement are

➢ The society believe a male partner should participate in earmarking of the skilled birth attendant,
➢ The society believe a male partner should escort his pregnant female partner during antenatal clinics,
➢ The society believe a male partner has to test for HIV with his pregnant female partner during antenatal visits,
➢ The society believe a male partner has to accompany his pregnant female partner during childbirth, and
➢ The society believe a male partner has to escort his female partner during postnatal checkups.

### Perceived behavior control is measured using the following Likert scale statements

➢ For me to participate in earmarking of the skilled birth attendant is with my pregnant woman is simple,
➢ For me to escort my pregnant woman during antenatal care clinics is simple,
➢ For me to test for HIV/AIDS with my pregnant woman during antenatal visits is simple,
➢ For me to accompany my female partner during labor and childbirth is simple, and
➢ For me to escort my female partner during postnatal checkups is simple.

### Validity and Reliability

As a tool was adopted from the study of exploring factors influencing pregnant Women’s attitudes, perceived subjective norms and perceived behavior control towards male involvement in maternal services utilization (Moshi et al., 2020). According to Moshi et al., (2020);

- A pilot study was undertaken to assess the accuracy of the data gathering tools in order to assure the tool’s validity.
- Cronbach’s Alpha was used to determine the tool’s dependability. Cronbach’s Alpha for attitude toward male involvement was 0.947, 0.948 for perceived subjective norms, and 0.938 for perceived behavior control.

### Variables description and measurement

#### Independent variables

In this study independent variables includes; male partners’ attitudes, perceived subjective norms, and perceived behavioral control, they influence male involvement in antenatal care clinics with their pregnant women.

#### Dependent variables

In this study male involvement in antenatal clinic in a dependent variable, it is influenced by male partners’ attitudes, perceived subjective norms, and perceived behavioral control which are independent variables.

### Data Analysis

The data were coded, checked, and processed with version 20 Statistical Package for the Social Sciences so as to generate descriptive statistical information which are then presented in form of tables, figures, pie-charts and bar graphs.

### Ethical Clearance

The ethical clearance committee of the university of Dodoma (UDOM), dean school of nursing and public health, granted authorization to perform the study. Permission was obtained from the municipal executive director (MED), and participants were given informed consent after being informed of the study’s benefits and objectives. Confidentiality was maintained throughout the study, and the questionnaire did not include the names of the respondents. Participants who agreed to take part in the study had the option to withdraw at any time without facing any penalty.

## Results

### Socio-demographic characteristics of sample

Out of 204 participants who were recruited as participant of the study by being thorough given and sign an informed consent elaborating the benefits and objectives of the study and that confidentiality, respect and dignity as well as participant’s autonomy was maintained throughout the study all consented to be a part of study making 100% response rate achieved in this study.

In regards to the age of the participants, majority of the respondents were aged 18 to 29 years (97, 47.5%), followed by age group 30 to 39 years (93, 45.6%) and lastly were aged group 40 to 49 years (14, 6.9%).

The majority of male partners were married (124, 60.8%), lived on more than $1 per day (183, 89.7%), and living in distance of less than 1km from a health facility (123, 60.3). 113, 55.4% of the responders have a secondary level of education. Larger number of participants reported that they are self-employed (89, 43.6%). In regards to religion most participants were Muslim (52.5%). In case of Health insurance (CHF, NHIF) possession majority of the respondents (147, 72.1%) reported that they do not possess health insurance (Table 1).

**Table 1:**
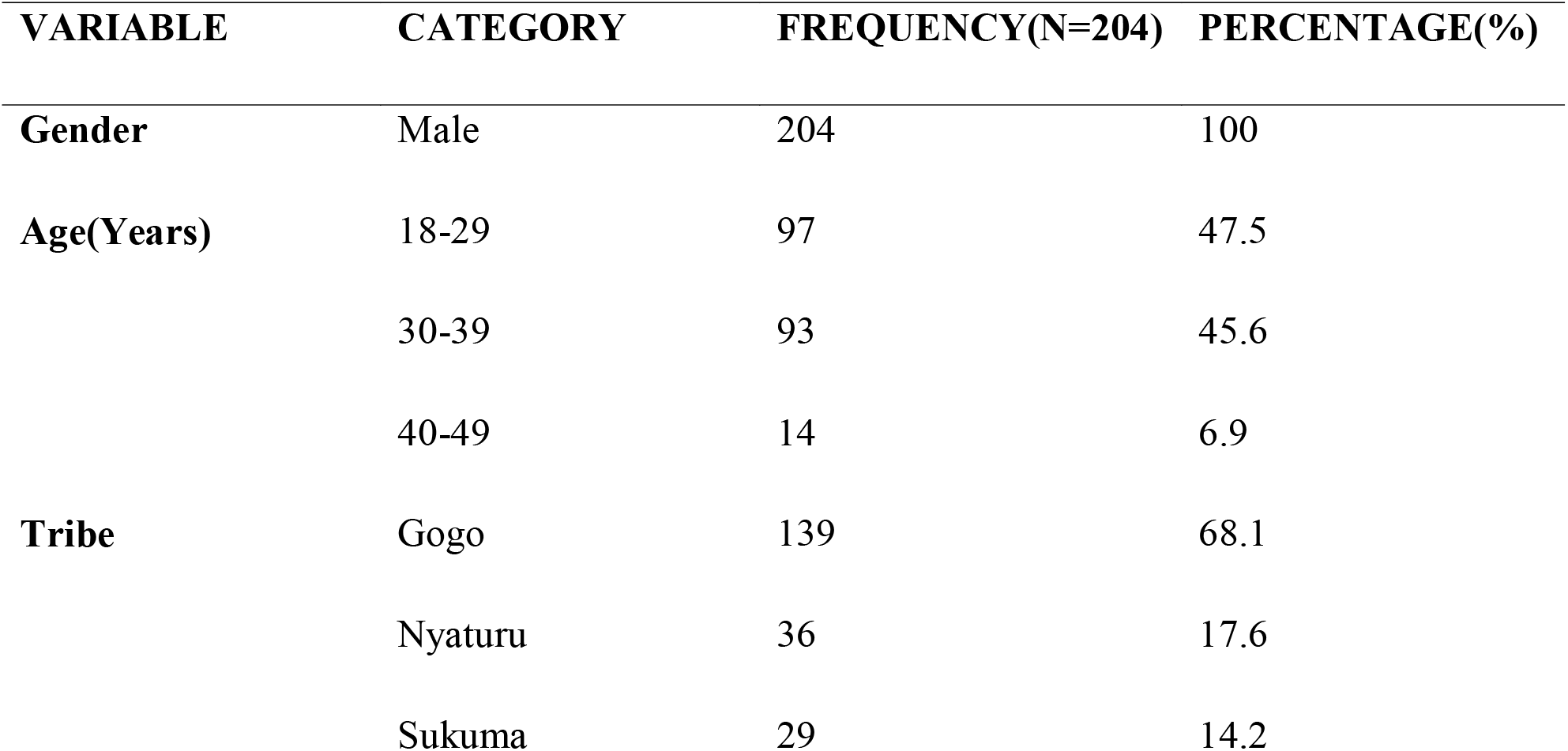
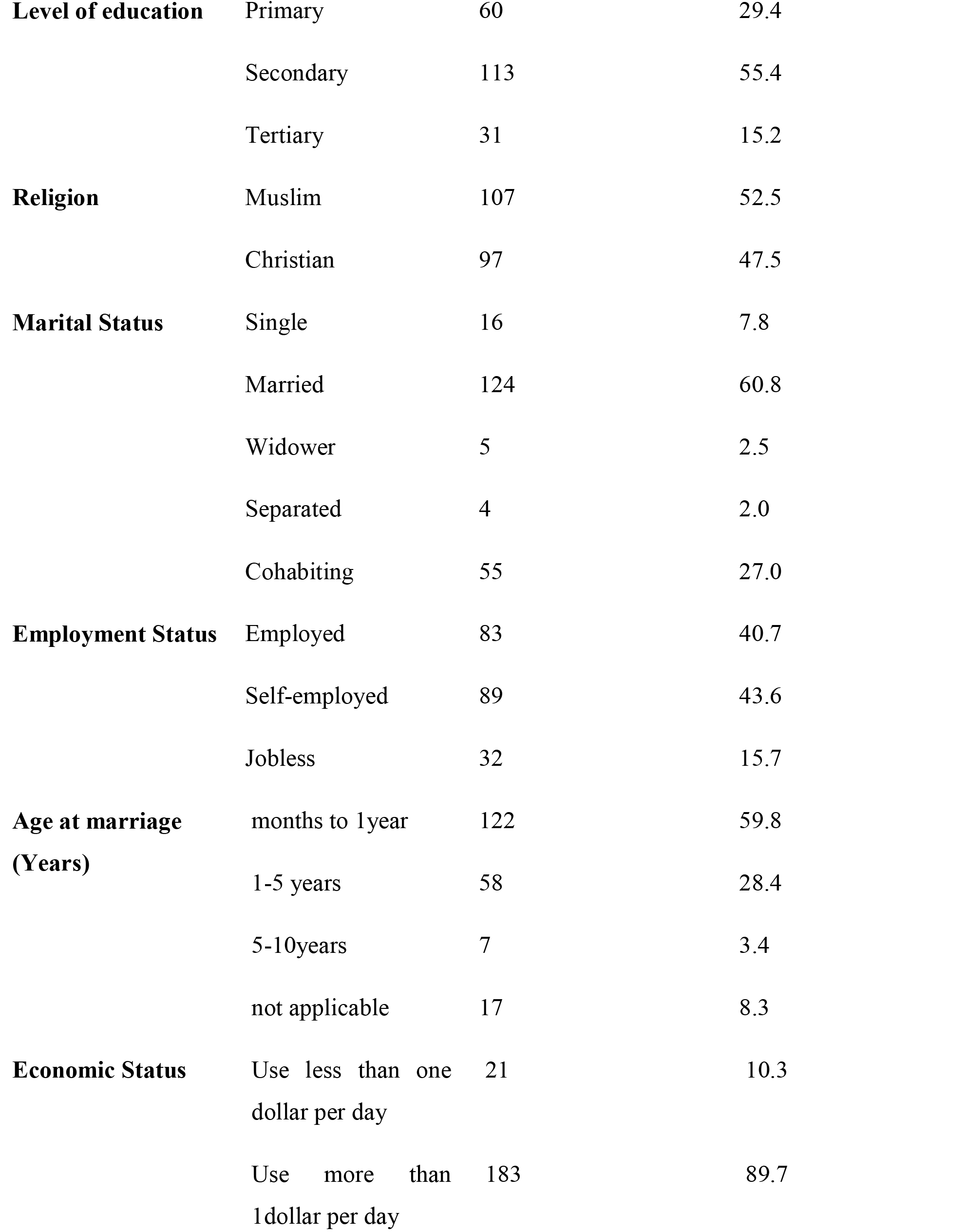
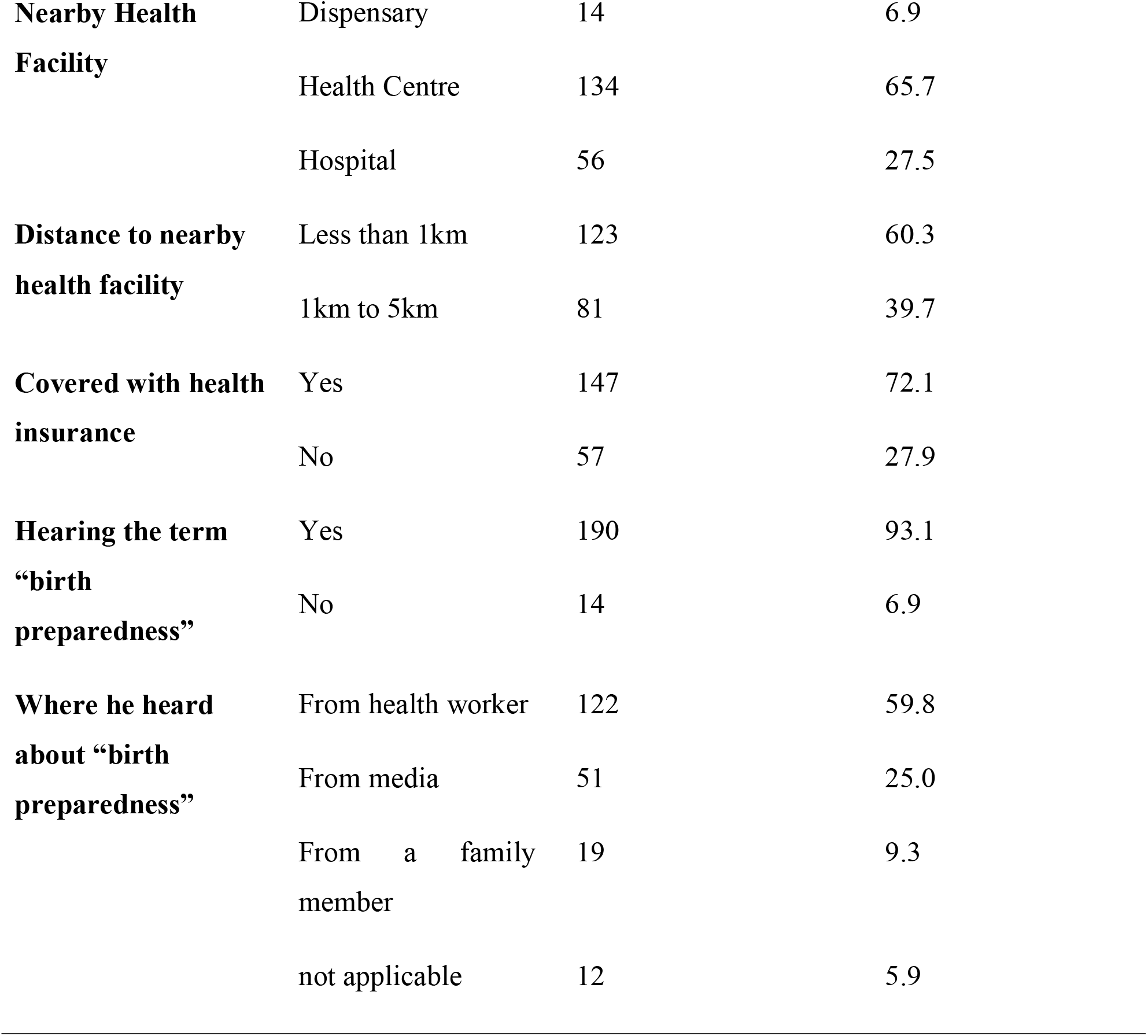
showing Socio-demographic characteristics of respondents.

| VARIABLE | CATEGORY | FREQUENCY(N=204) | PERCENTAGE(%) |
| --- | --- | --- | --- |
| **Gender** | Male | 204 | 100 |
| **Age(Years)** | 18-29 | 97 | 47.5 |
|  | 30-39 | 93 | 45.6 |
|  | 40-49 | 14 | 6.9 |
| **Tribe** | Gogo | 139 | 68.1 |
|  | Nyaturu | 36 | 17.6 |
|  | Sukuma | 29 | 14.2 |
| **Level of education** | Primary | 60 | 29.4 |
|  | Secondary | 113 | 55.4 |
|  | Tertiary | 31 | 15.2 |
| **Religion** | Muslim | 107 | 52.5 |
|  | Christian | 97 | 47.5 |
| **Marital Status** | Single | 16 | 7.8 |
|  | Married | 124 | 60.8 |
|  | Widower | 5 | 2.5 |
|  | Separated | 4 | 2.0 |
|  | Cohabiting | 55 | 27.0 |
| **Employment Status** | Employed | 83 | 40.7 |
|  | Self-employed | 89 | 43.6 |
|  | Jobless | 32 | 15.7 |
| **Age at marriage**<br>**(Years)** | months to 1year | 122 | 59.8 |
|  | 1-5 years | 58 | 28.4 |
|  | 5-10years | 7 | 3.4 |
|  | not applicable | 17 | 8.3 |
| **Economic Status** | Use less than one<br>dollar per day | 21 | 10.3 |
|  | Use more than<br>1dollar per day | 183 | 89.7 |
| **Nearby Health Facility** | Dispensary | 14 | 6.9 |
|  | Health Centre | 134 | 65.7 |
|  | Hospital | 56 | 27.5 |
| **Distance to nearby health facility** | Less than 1km | 123 | 60.3 |
|  | 1km to 5km | 81 | 39.7 |
| **Covered with health insurance** | Yes | 147 | 72.1 |
|  | No | 57 | 27.9 |
| **Hearing the term “birth preparedness”** | Yes | 190 | 93.1 |
|  | No | 14 | 6.9 |
| **Where he heard about “birth preparedness”** | From health worker | 122 | 59.8 |
|  | From media | 51 | 25.0 |
|  | From a family member | 19 | 9.3 |
|  | not applicable | 12 | 5.9 |

**Table 2:** Frequency table.

| **Male's Attitude towards their involvement in ANC visits** | **Frequency** | **Percent (%)** |
| --- | --- | --- |
| **Negative(6 to 23)** | 21 | 10.3 |
| **Positive(24 to 30)** | 183 | 89.7 |
| **Total** | 204 | 100.0 |
| **Male's perceived subjective norms towards their involvement in ANC visits** | Frequency | Percent (%) |
| **Negative (8 to 31)** | 44 | 21.6 |
| **Positive (32 to 40)** | 160 | 78.4 |
| **Total** | 204 | 100.0 |
| **Male's perceived behavior control towards their involvement in ANC visits** | Frequency | Percent (%) |
| **Negative (8 to 31)** | 41 | 20.1 |
| **Positive (32 to 40)** | 163 | 79.9 |
| **Total** | 204 | 100.0 |

Male partners’ attitudes, perceived subjective norms and perceived behavior control towards their involvement in ANC with their pregnant women at Dodoma

By considering table 2 above, concerning attitude, (183,89.7%) of respondents (male partners) have a positive attitude, while the remaining (21,10.3%) of the respondents have a negative attitude towards their involvement in ANC visits with their pregnant women (Table).

On the other hand, perceived subjective norms, (160,78.4%) of respondents (male partners) have positive perceived subjective norms, while the remaining (44,21.6%) of the respondents have a negative perceived subjective norms towards their involvement in ANC visits with their pregnant women (Table 2).

Furthermore, perceived behavior control, (163,79.9%) of respondents (male partners) have positive perceived behavior control, while the remaining (41,20.1%) of the respondents have a negative perceived behavior control towards their involvement in ANC visits with their pregnant women (Table 2).

**Association of socio-demographic characteristics and male partners’ attitude, perceived subjective norms and perceived behavior control towards their involvement in ANC with their pregnant women (Chi-squire Test).**

**Table 3:**
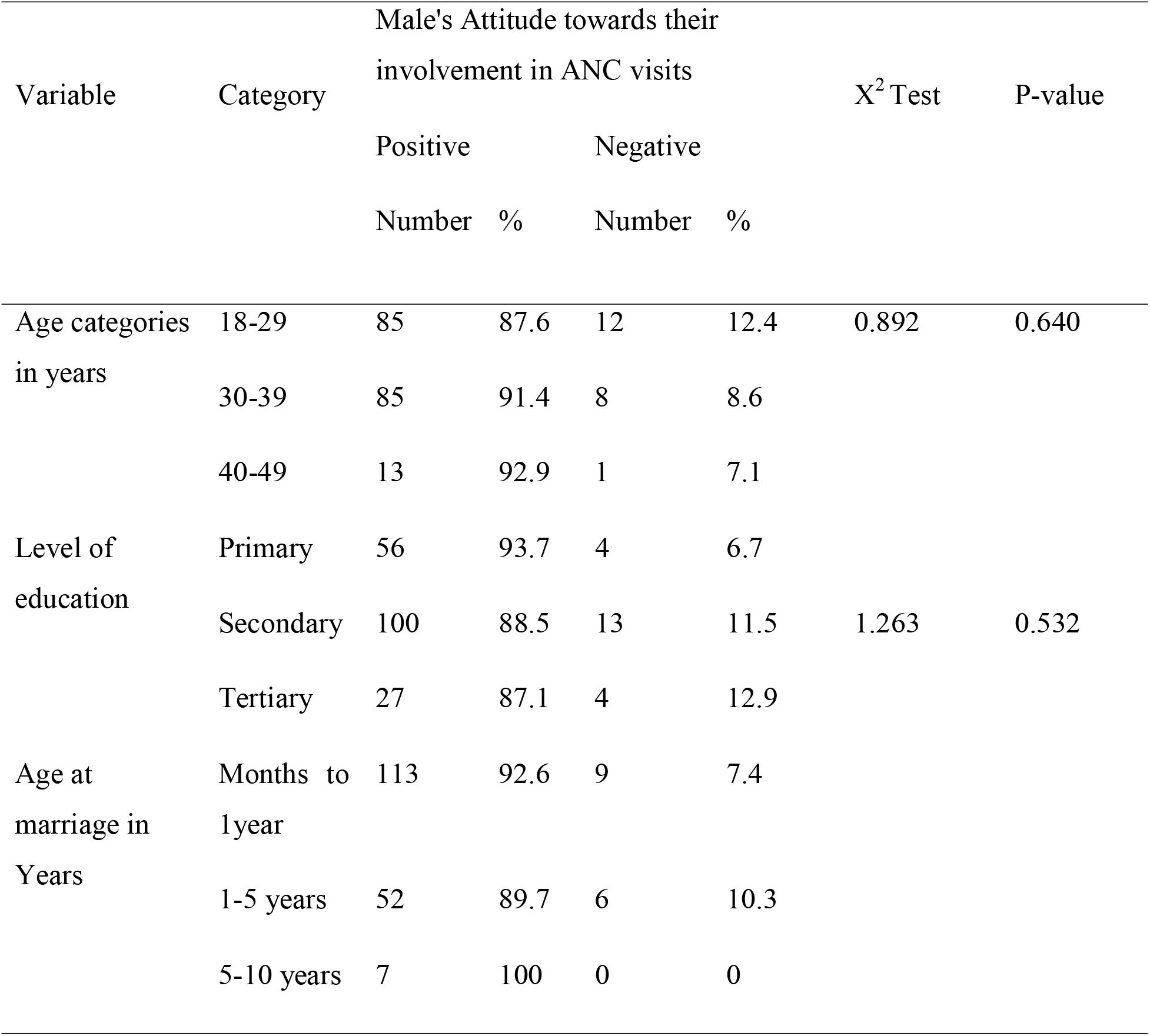
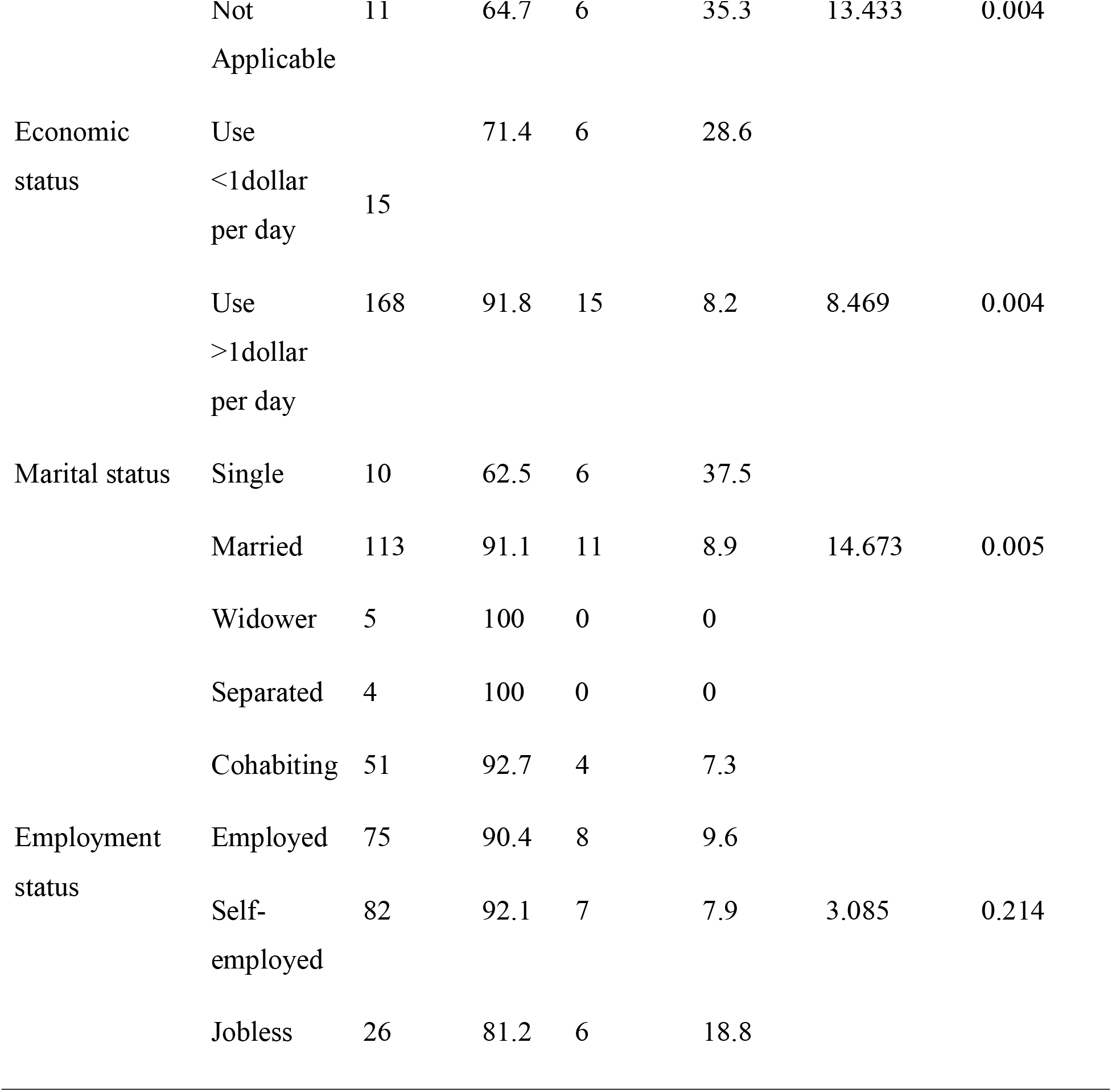
Association of socio-demographic characteristics and male partners’ attitude towards their involvement in ANC with their pregnant women.

From the Table 3 above, the variables which their relationship with male partners’ attitudes towards their involvement in ANC visits with their pregnant women were statistically significant were economic status (p < 0.05), age at marriage (p < 0.05) and marital status (p < 0.05) (Table 3)

**Table 4:**
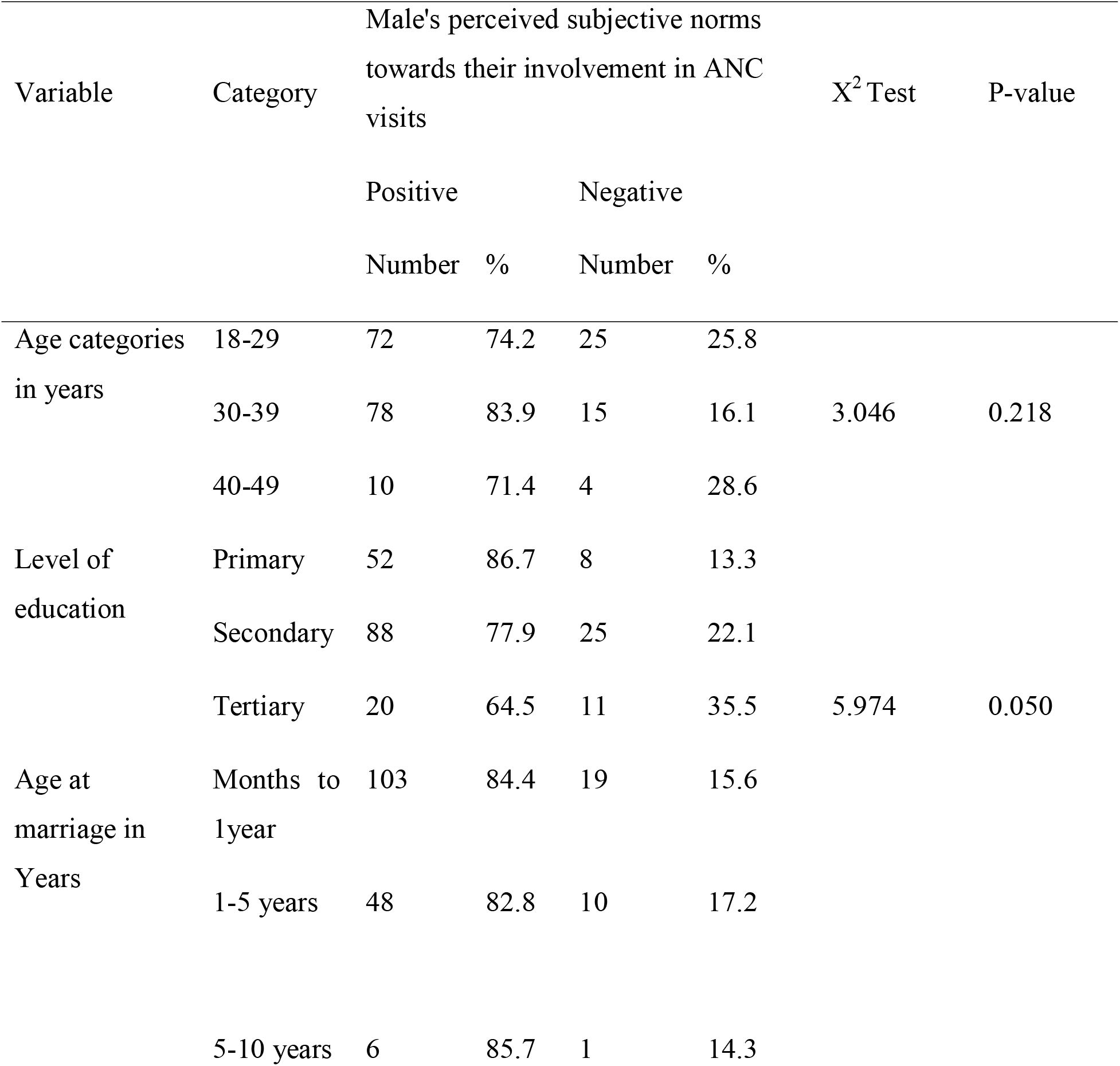
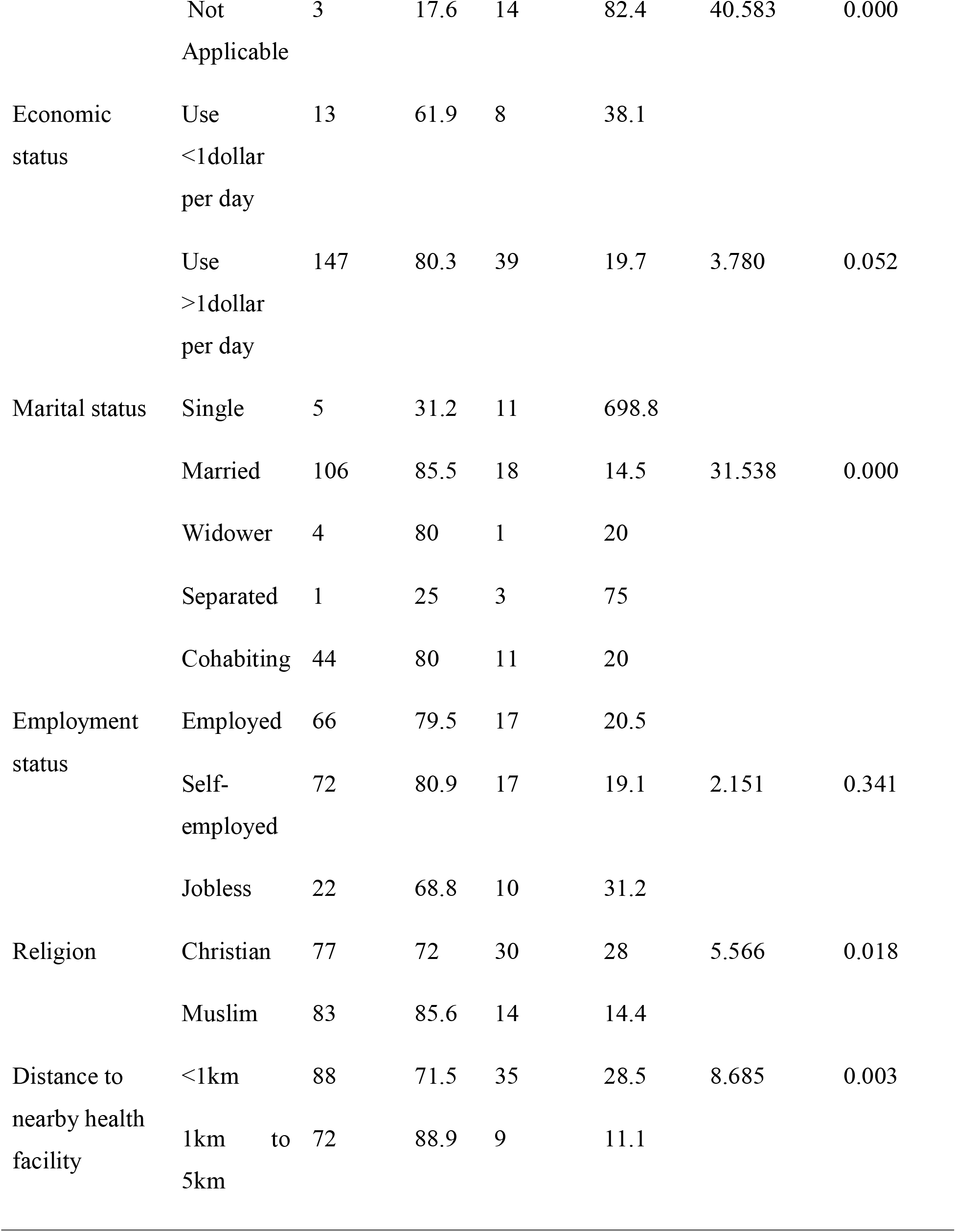
Association of socio-demographic characteristics and male partners’ perceived subjective norms towards their involvement in ANC with their pregnant women.

From the Table 4 above, the variables which their relationship with male partners’ perceived subjective norms towards their involvement in ANC visits with their pregnant women is statistically significant were level of education (p=0.05), age at marriage (p < 0.001), marital status (p < 0.001), religion (p<0.05) and distance to nearby health facility (p<0.05) (Table 4).

**Table 5:**
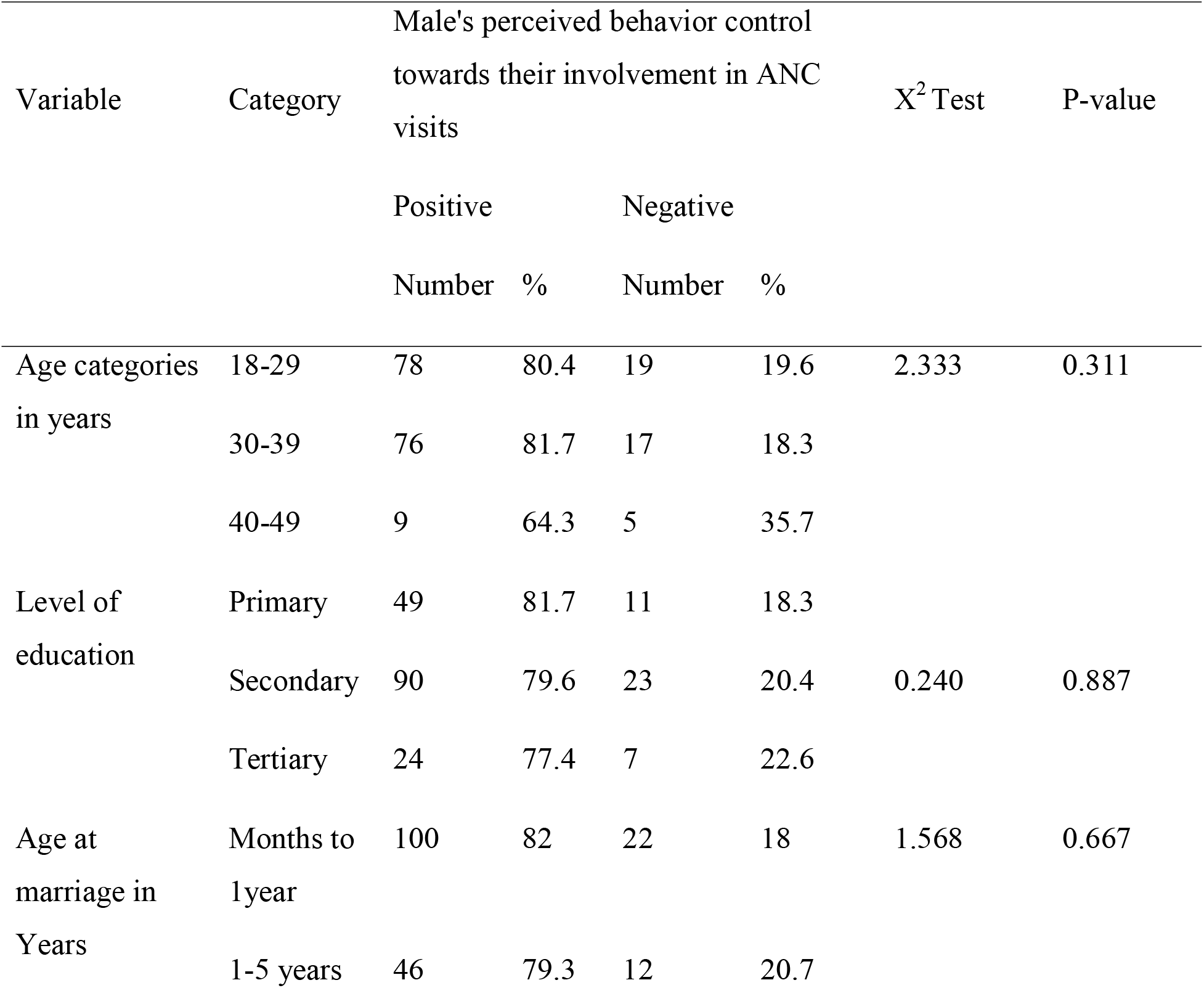
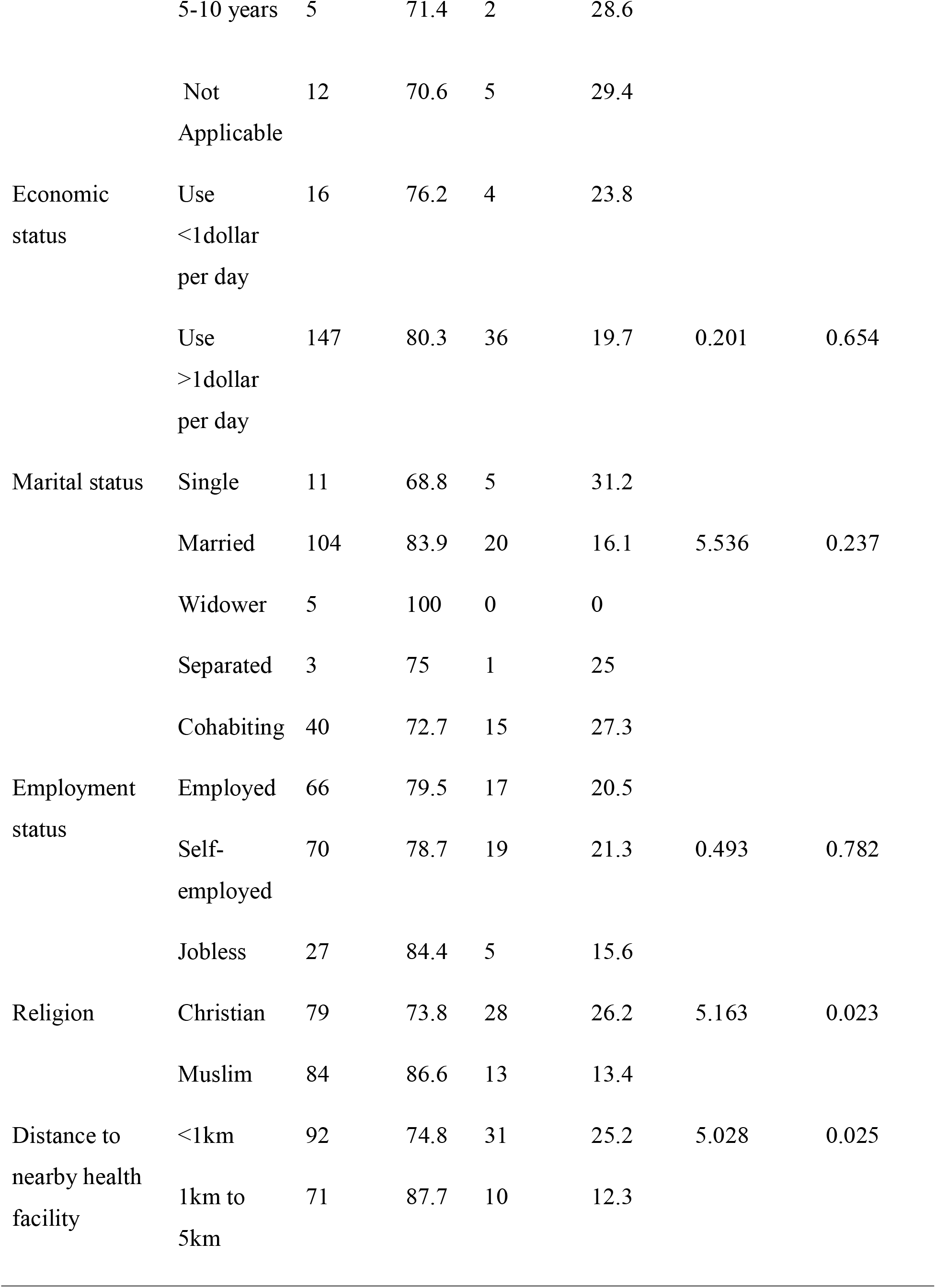
Association of socio-demographic characteristics and male partners’ perceived behavior control towards their involvement in ANC with their pregnant women.

From the Table 5 above, the variables which their relationship with male partners’ perceived behavior control towards their involvement in ANC visits with their pregnant women is statistically significant were religion (p<0.05) and distance to nearby health facility (p<0.05) (Table 5).

**Association of socio-demographic characteristics and male partners’ attitude, perceived subjective norms and perceived behavior control towards their involvement in ANC with their pregnant women (Logistic Regression Analysis).**

**Table 6:**
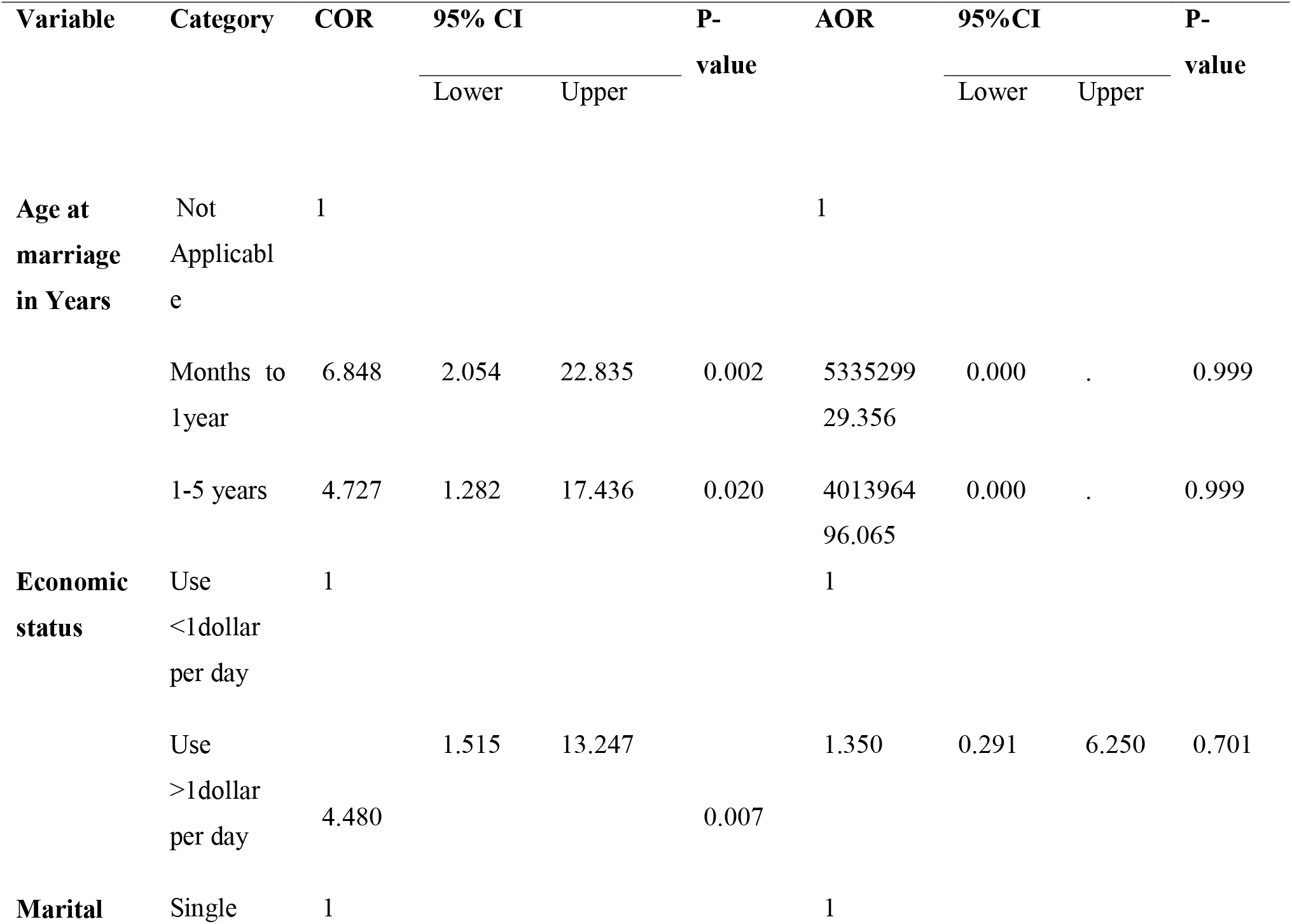
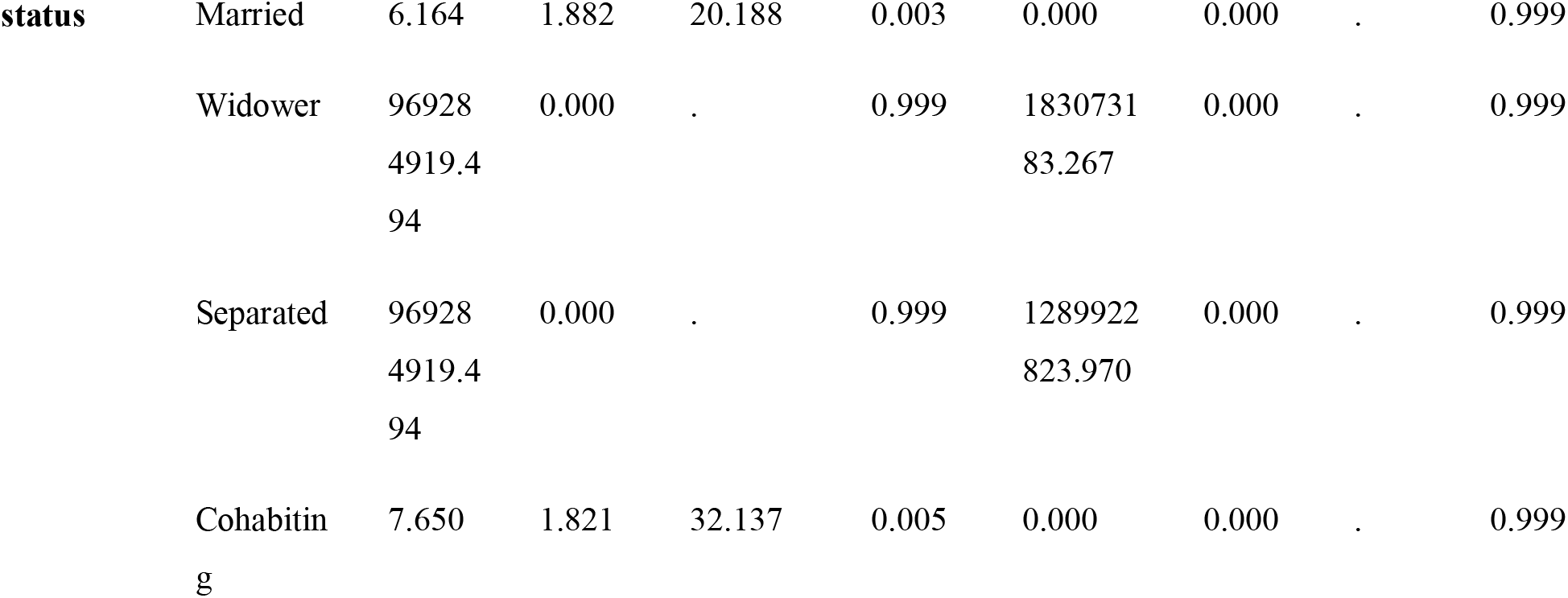
Association between socio-demographic factors with Male’s Attitude towards their involvement in ANC visits (Logistic regression analysis)

From Table 6 above, Binary logistic regression was used to assess the association between socio-demographic factors with Male’s Attitude towards their involvement in ANC visits and the following are the factors involved:

### On crudes odds ratio (COR) the findings were as follows

In regarding to Age at marriage in years (COR: 4.727, 95%CI: 1.282 -17.436, P= 0.020) was associated with Male’s Attitude towards their involvement in ANC visits implying a unit increase in age at marriage increases with individual Male’s Attitude towards their involvement in ANC visits and its association is statistical significant. Hence the male partners age group at marriage 1-5 years is 4.727 more likely to get involved in ANC visits with their pregnant women as compared to other groups (Table 6).

In regarding to economic status (COR: 4.480, 95%CI:1.515-13.247, P= 0.007), male partners who Use more than 1dollar per day are 4.480 more likely to get involved in ANC visits with their pregnant women as compared to those who use less than 1dollar per day (Table 6).

In regarding to Marital Status (COR: 6.164, 95%CI: 1.882-20.188, P= 0.003), married male partners were 4.480 more likely to get involved in ANC visits with their pregnant women as compared to other groups (Table 6).

#### On Adjusted Odds Ratio (AOR)

The findings show that, there is no association between socio-demographic factors with Male’s Attitude towards their involvement in ANC visits as in all socio-demographic factors p-value>0.05.

From Table 7 above, Binary logistic regression was used to assess the association between socio-demographic factors with Male’s perceived subjective norms towards their involvement in ANC visits and the following are the factors involved:

**Table 7:**
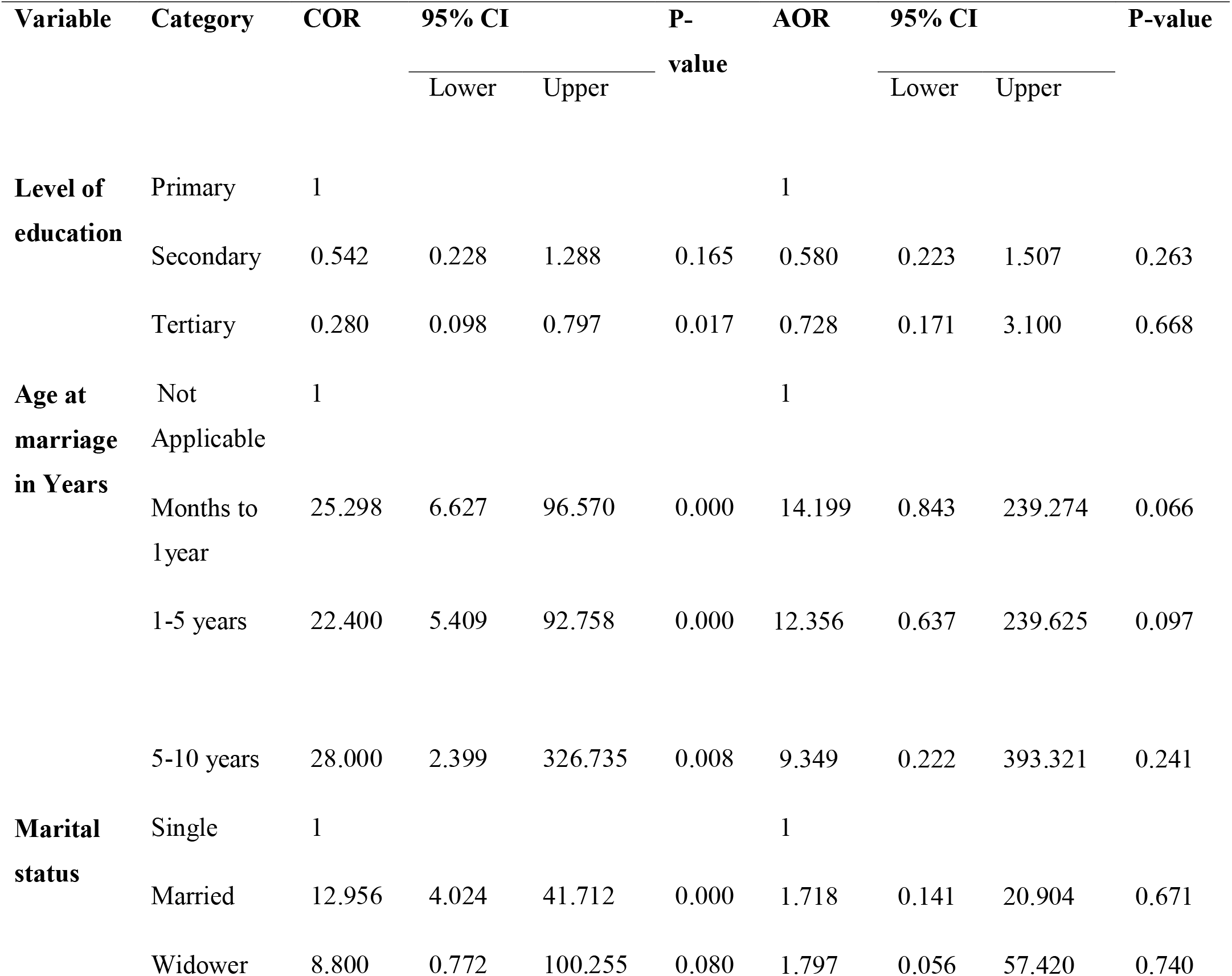
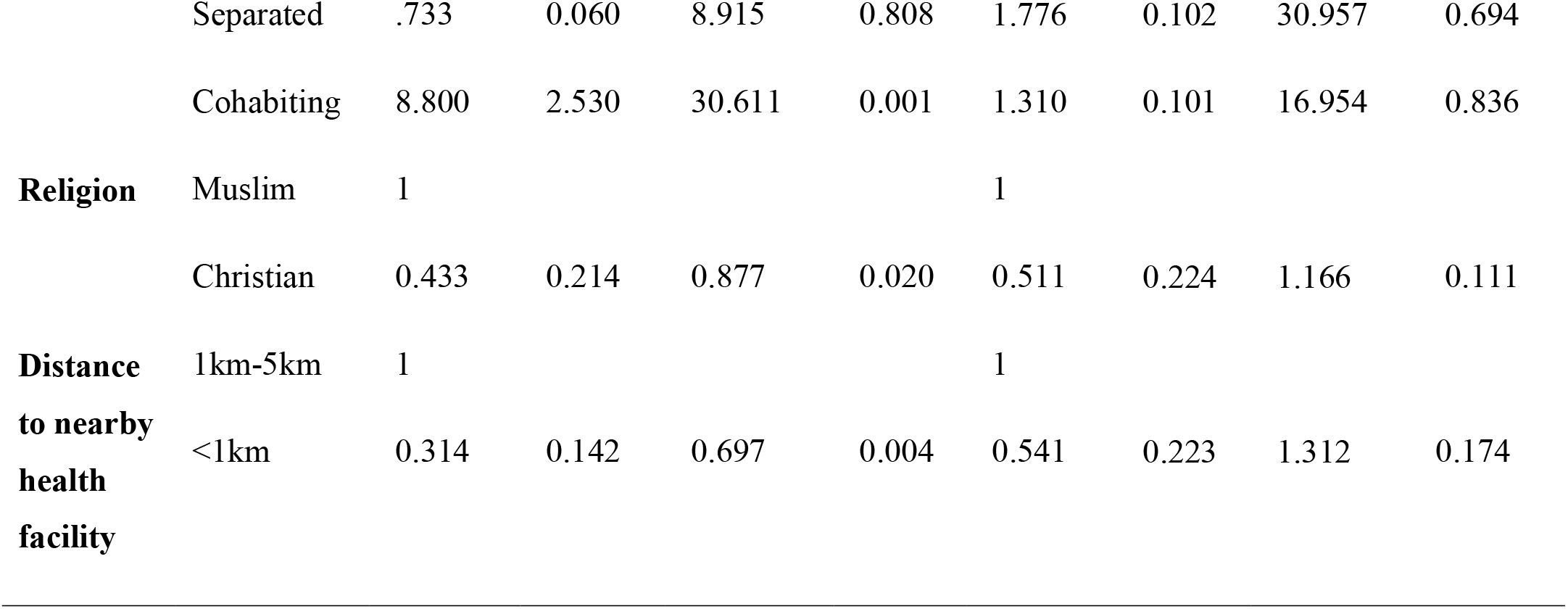
Association between socio-demographic factors with Male’s perceived subjective norms towards their involvement in ANC visits (Logistic regression analysis).

### On crudes odds ratio (COR) the findings were as follows

In regarding to Level of education (COR: 0.280, 95%CI: 0.098-0.797, P= 0.017), unit increase in level of education increases with individual Male’s perceived subjective norms towards their involvement in ANC visits and its association is statistical significant. Hence the male partners tertiary level of education (colleges, universities) is 0.280 more likely to get involved in ANC visits with their pregnant women as compared to other levels (Table 7).

In regarding to Age at marriage in years (COR: 28.000, 95%CI: 2.399 -326.735, P= 0.008) was associated with Male’s perceived subjective norms towards their involvement in ANC visits implying a unit increase in age at marriage increases with individual Male’s perceived subjective norms towards their involvement in ANC visits and its association is statistical significant. Hence the male partners age group at marriage 5-10 years is 28.000 more likely to get involved in ANC visits with their pregnant women as compared to other groups (Table 7).

In regarding to Marital Status (COR: 12.956, 95%CI: 4.024-41.712, P<0.001), married male partners were 12.956 more likely to get involved in ANC visits with their pregnant women as compared to other (Table 7).

In regarding to distance to nearby health facility (COR: 0.314, 95%CI: 0.142-0.697, P= 0.004), male partners who live in a distance less than 1 kilometer from the nearby health facility are 0.314 more likely to get involved in ANC visits with their pregnant women as compared to those who live in a distance more than 1 kilometer from the nearby health facility (Table 7).

In regarding to Religion (COR: 0.433, 95%CI: 0.214-0.877, P=0.020), Christian male partners were 0.433 more likely to get involved in ANC visits with their pregnant women as compared to other religions (Table 7).

#### On Adjusted Odds Ratio (AOR)

The findings show that, there is no association between socio-demographic factors with Male’s perceived subjective norms towards their involvement in ANC visits as in all socio-demographic factors p-value>0.05 (Table 7).

From Table 8 above, Binary logistic regression was used to assess the association between socio-demographic factors with Male’s perceived behavior control towards their involvement in ANC visits and the following are the factors involved:

**Table 8:**
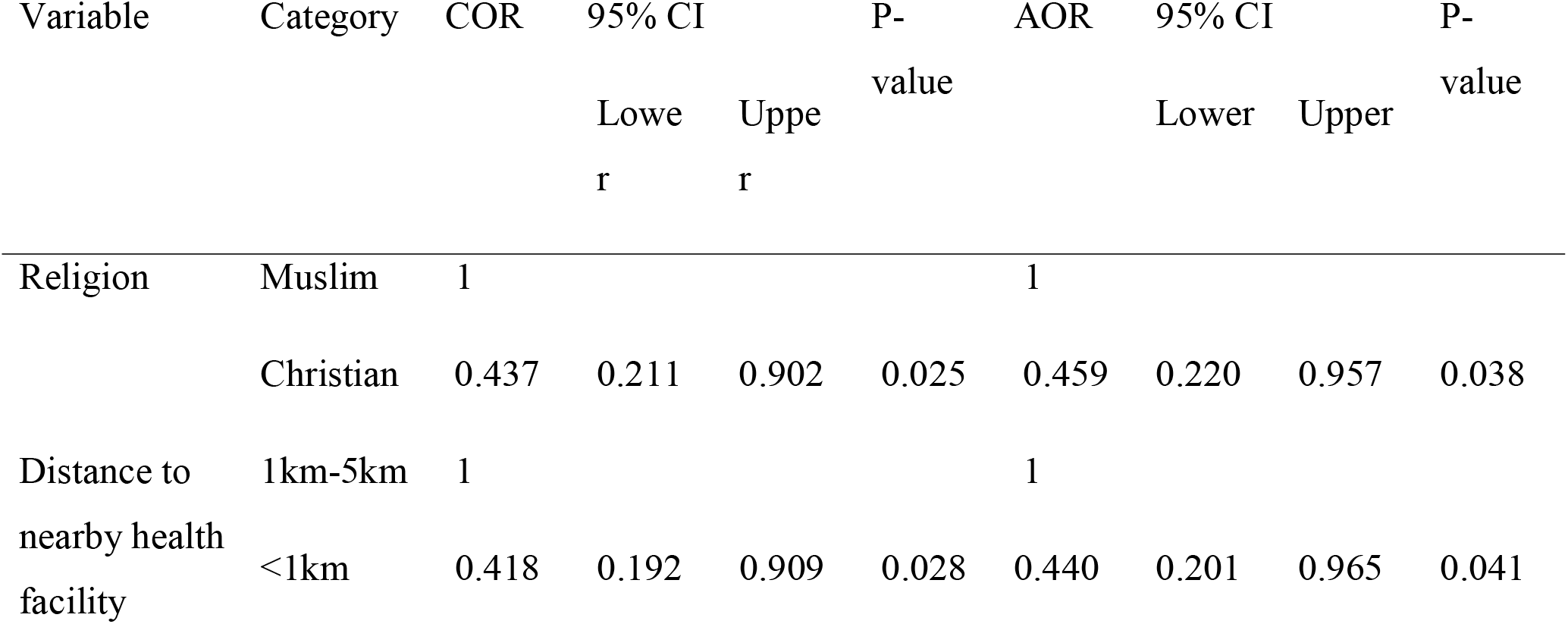
Association between socio-demographic factors with Male’s perceived behavior control towards their involvement in ANC visits (Logistic regression analysis)

### On crudes odds ratio (COR) the findings were as follows

In regarding to Religion (COR: 0.437, 95%CI: 0.211-0.902, P=0.025), Christian male partners were 0.437 more likely to get involved in ANC visits with their pregnant women as compared to other religions (Table 8).

In regarding to distance to nearby health facility (COR: 0.418, 95%CI: 0.192-0.909, P= 0.028), male partners who live in a distance less than 1 kilometer from the nearby health facility are 0.418 more likely to get involved in ANC visits with their pregnant women as compared to those who live in a distance more than 1 kilometer from the nearby health facility (Table 8).

### On Adjusted Odds Ratio (AOR)

After adjusting the cofounders, the findings show that there was statistical significant association between religion and distance from the nearby health facility with male partners’ perceived behavior control towards their involvement in ANC visits, as their p-value<0.05 (Table 8).

## Discussions

The majority of maternal deaths occur during birth and the immediate postpartum period, which can be avoided by eliminating three delays during an emergency, namely the time it takes to seek, reach, and receive medical assistance. Males, who are typically the decision-makers, can help to eliminate these delays and thereby improve pregnancy outcomes by participating actively in antenatal care clinics (Paper et al., 2019).

### Individual beliefs and assessments of behavioral results have an impact on attitude

The way a male partner feels his society approves or disapproves of male engagement in antenatal visits with their pregnant woman is what is referred to as perceived subjective norms. He will act in favor of societal approval if he believes his society accepts of him accompanying his female companion, but he will act accordingly if he believes his society disapproves of the behavior.

As a result, it was necessary to evaluate the attitudes, subjective norms, and behavior of male partner’s engagement in antenatal care visits with their pregnant women is believed to be under control (Moshi et al., 2020).

From this study finding it was revealed that majority of male partners have positive attitudes, perceived subjective norms and perceived behavior control; -

#### Male partners’ attitudes

On attitude, (183,89.7%) of male partners have positive attitude, while the remaining (21,10.3%) have negative attitude towards their involvement in ANC visits with their pregnant women (Table2). Male partners’ attitudes towards their involvement in ANC visits with their pregnant women were influenced by male partners’ economic status where large number with positive attitudes were those using more than one dollar per day, age at marriage where an increase in age at marriage goes with positive attitudes and marital status where large number of married male partners have positive attitudes compared to others. In comparison to men who live apart from their wives, men who are married and live together are more likely to participate in antenatal care with their pregnant women because they have more time to spend with their partners and teach them about the value of male involvement (Factors & Facility, 2013).

#### Male partners’ perceived subjective norms

On perceived subjective norms, (160,78.4%) male partners have positive perceived subjective norms, while the remaining (44,21.6%) have negative perceived subjective norms towards their involvement in ANC visits with their pregnant women (Table2). Male partners’ perceived subjective norms towards their involvement in ANC visits with their pregnant women were influenced by level of education those with tertiary level (colleges and universities) have positive perceived subjective norms compared to others, those who have never attended school, on the other hand, have limited access to health-related information (Chibwae et al., 2018). age at marriage, marital status large number of married had positive, religion and distance to nearby health facility where large number of those living less than 1kilometer away from health facility had positive perceived subjective norms. closeness to the health center plays a role in male participation because those who live close to the health center are more likely to participate because it will take them less time to get there than those who live further away (Gibore et al., 2019).

#### Male partners’ perceived behavior control

And on perceived behavior control, (163,79.9%) male partners have positive perceived behavior control, while the remaining (41,20.1%) have negative perceived behavior control towards their involvement in ANC visits with their pregnant women (Table2). Male partners’ perceived behavior control towards their involvement in ANC visits with their pregnant women were influenced by religion and distance to nearby health facility, large number of those living less than 1kilometer away from health facility had positive perceived subjective norms.

## Limitations of study

The study has limitations because only male partners were interviewed. Interviews with female partners may reveal new or different information about their thoughts on attitudes, perceived subjective norms and perceived behavior control on men Involvement in ANC clinics with their pregnant women. Additionally, because this study was only conducted in one district, the results could not accurately represent what other Tanzanian districts have gone through.

## Conclusions

From this study, Male partners’ attitudes, perceived subjective norms and perceived behavior control towards their involvement in ANC visits with their pregnant women were influenced by level of education those with tertiary level (colleges and universities) have positive perceived subjective norms compared to others, economic status where large number with positive attitudes were those using more than one dollar per day, age at marriage where an increase in age at marriage goes with positive attitudes and marital status where large number of married male partners have positive attitudes compared to others. So interventions must be done in those areas in order to reduce maternal complications hence pregnant women’s health.

## Recommendation

As the study found majority of male partners had positive attitudes, perceived subjective norms and perceived behavior control towards their involvement in ANC with their women, so we must strengthen those factors associated with male partners’ attitudes, perceived subjective norms and perceived behavior control for better outcomes on maternal and reproductive health in general.

This study has recommendation to stakeholders in maternity health care and reproductive health, based on the results from this study stakeholder who are program planners too should redesign maternity care program in which will regard men as key player in the provision of maternity care to pregnant women for better outcomes. This will provide awareness to both men and women hence improve participation and maternity outcome.

## Declaration

Authors declare no competing interest.

## Supporting information

https://us.docworkspace.com/d/sIG6_9Z0isba0sAY

## Data Availability

All data produced in the present study are available upon reasonable request to the authors

## Acknowledgement

We are extremely grateful to God for his guidance, protection and granting wellbeing during the preparation of this work. This work would not have been possible without the guidance of the University of Dodoma, specifically school of nursing and public health lecturers. Special regards and appreciation to Dr. Fabiola V. Moshi, our supervisor for her intelligent, guidance, encouragement, support and patience.

## References

Ajzen, I. (2014). Attitudes and the Attitude-Behavior Relation_J: Reasoned and Automatic Processes. January 2000. 10.1080/14792779943000116

Alemi, S., Nakamura, K., Rahman, M., & Seino, K. (2020). *Male participation in antenatal care and its influence on their pregnant partners ’ reproductive health care utilization*_J*: insight from the 2015 Afghanistan Demographic and Health Survey*. 10.1017/S0021932020000292

Annoon, Y., Hormenu, T., Opoku, B., Seidu, A., Kwabena, E., & Sambah, F. (2020). Heliyon Perception of pregnant women on barriers to male involvement in antenatal care in Sekondi , Ghana. Heliyon, 6(November 2019), e04434. 10.1016/j.heliyon.2020.e04434

Chibwae, A., Kapesa, A., Jahanpour, O., Seni, J., Namanya, B., Kadelya, E., Konje, E., Nyanza, E. C., Ngallaba, S., & Dewey, D. (2018). Attendance of male partners to different reproductive health services in Shinyanga district, north western Tanzania. Tanzania Journal of Health Research, 20(2), 1–11. 10.4314/thrb.v20i2.9

Factors, R., & Facility, H. (2013). original ARTICLE. 7(3), 46–54.

Gibore, N. S., Bali, T. A. L., & Kibusi, S. M. (2019). *Factors influencing men ’ s involvement in antenatal care services*_J*: a cross-sectional study in a low resource setting* , Central Tanzania. 1–10.

Kabanga, E., Chibwae, A., Basinda, N., & Morona, D. (2019). Prevalence of male partners involvement in antenatal care visits – in Kyela district , Mbeya. 4, 1–6.

Messages, K., Recommendations, G., & Care, R. A. (2018). WHO Recommendations on Antenatal Care for a Positive Pregnancy Experience_J: Summary Highlights and Key Messages from the World Health Organization ’ s 2016 Global Recommendations for Routine Antenatal Care. 10(January), 1–10. 10.1186/1742-4755-10-19.5

Mgata, S., & Maluka, S. O. (2019). Factors for late initiation of antenatal care in Dar es Salaam , Tanzania_J: A qualitative study. 0, 1–9.

Moshi, F. V, Kibusi, S. M., & Fabian, F. (2020). Exploring factors influencing pregnant Women ’ s attitudes , perceived subjective norms and perceived behavior control towards male involvement in maternal services utilization_J: a baseline findings from a community based interventional study from Rukwa , rural Tanzania. 1–12.

Natai, C. C., Gervas, N., Sikira, F. M., Leyaro, B. J., Mfanga, J., Yussuf, M. H., & Msuya, S. E. (2020). Association between male involvement during antenatal care and use of maternal health services in Mwanza City , Northwestern Tanzania_J: a cross- - sectional study. 10.1136/bmjopen-2019-036211

Nesbitt, U. C. A. R. C. (2017). UNICEF ’ s application of the new ANC recommendations_J: Actions to reduce the burden of Malaria in Pregnancy Valentina Buj , Global Malaria Advisor. September.

Paper, O., Beksac, A. T., Orgul, G., Tanacan, A., Uckan, H., Sancak, B., Portakal, O., & Beksac, M. S. (2019). Uropathogens and Gestational Outcomes of Urinary Tract Infections in Pregnancies that Necessitate Hospitalization. 70–73. 10.1159/000499290

Republic, T. U., Bureau, N., Ministry, S., & June, F. (2013). TANZANIA IN FIGURES 2012.

