## Supplementary material for "Assessing Male Partners’ Attitudes, Perceived Subjective Norms and Perceived Behavior Control Towards Their Involvement in Antenatal Care Clinic with Their Pregnant Women in Dodoma Urban Municipal": https://us.docworkspace.com/d/sIG6_9Z0isba0sAY

### Questionnaire in English and Swahili format

**English Version Questionnaires**

**First questionnaire: Male Partners**

**Part A: Baseline Information**

**Please tick (√) the appropriate option**

1. Age in years in years______________

2. Age at marriage in years_________

3. Phone number__________________

4. Marital status
   a) Married ( )
   b) Cohabited ( )

5. Educational status
   a) None ( )
   b) Primary level incomplete ( )
   c) Primary level complete ( )
   d) Secondary or higher ( )

6. Employment status
   a) Employed ( )
   b) Unemployed ( )

7. Religion
   a) Christian ( )
   b) Muslims ( )
   c) Others specify………………………

8. Ethnic group
   a) Nyaturu ( )
   b) Sukuma ( )
   c) Others,specify………………………

9. Economic status
   a) Use less than one dollar per day ( )
   b) Use more than one dollar per day ( )

10. Do you own radio?
    a) Yes ( )
    b) No ( )

11. Do you own mobile phone?
    a) Yes ( )
    b) No ( )

12. Characteristic of a nearby health facility
    a) Dispensary ( )
    b) Health center ( )
    c) Hospital ( )

13. What is the walking distance to a nearby health facility?
    a) Less than one kilometer ( )
    b) One kilometer-5kilometers ( )
    c) More than five kilometers ( )

14. Are you covered with health Insurance (NHIF or CHF)?
    a) Yes ( )
    b) No ( )
15. Have you ever heard the term “birth preparedness”?
    a) Yes ( ) b) No ( )
16. Where did you hear about birth preparedness?
    a) From health worker ( )
    b) From the media ( )
    c) From a family member ( )
    d) Others, specify…………

**Part B: Male partners’ behavior intentions towards their involvement in antenatal care visits with their pregnant women**

Many questions in this survey make use of rating scales with 5 places; you are supposed to tick (√) the box that best describes your opinion where SA= strongly agree, A= agree, N= Neutral, D= disagree and SD = strongly disagree

| Component | SA(5) | A (4) | N (3) | D (2) | SD(1) |
|---|---|---|---|---|---|
| **B: Male involvement** | | | | | |
| ***Attitude of male partners towards male involvement*** | | | | | |
| 1. If male partner participates in setting a part of the skilled birth attendant, he is doing a good and beneficial thing, | | | | | |

| |
|---|
| 2. If a male partner accompanies his pregnant female partner during antenatal clinics, he is doing a good and beneficial thing, |
| 3. If male partner tests for HIV with his female partner during pregnancy, he is doing a good and beneficial thing, |
| 4. If male partner accompanies his female partner during childbirth, he is doing a good thing which is beneficial, |
| 5. If male partner accompanies his female partner for postnatal checkups, he is doing a good and beneficial thing |
| 6. If a male partner test for syphilis with his pregnant female partner during antenatal clinics he is doing a good thing which is beneficial |
| ***Assessment of subjective norms*** |
| 7. The society think a male partner has to participate in setting aside funds and equipments to be used in case of emergency or during childbirth. |
| 8. The society think a male partner has to participate in preparation of transport to be used in case of emergency or during childbirth |
| 9. The society think a male partner should participate in identification of skilled birth attendant |
| 10. The society think a male partner has to accompany his pregnant female partner during antenatal clinics |
| 11. The society think a male partner has to test for HIV with his pregnant female partner during antenatal visits |

| |
|---|
| 12. The society think that a male partner has to screen for syphilis with his pregnant female partner during antenatal visit |
| 13. The society think that a male partner has to accompany his pregnant female partner during childbirth |
| 14. The society think that a male partner has to accompany his female partner during postnatal checkups |
| ***Perceived Behavioral Control*** |
| 15. For you to participate in setting aside funds and equipments to be used in case of emergency or during childbirth is simple and you can do it |
| 16. For you to participate in preparation of transport to be used in case of emergency or during childbirth is simple and you can do it |
| 17. For you to participate in identification of skilled birth attendant is simple and you can do it |
| 18. For you to accompany pregnant female partner during antenatal clinics is simple and you can do it |
| 19. For you to test for HIV/AIDS with your pregnant female partner during antenatal visits is trouble free and you can do it |
| 20. For you to test for syphilis with your female partner during antenatal clinics is simple and you can do it |
| 21. For you to accompany your pregnant female partner during labor and childbirth is simple and you can do it. |
| 22. For you to accompany your female partner during postnatal checkup is trouble free and you can do it |

**Thank You For Accepting To Be Part Of This Study.**

**Swahili version questionnaire**

**Hojaji ya kwanza: Washirika wa Kiume**

**Sehemu A: Taarifa za Msingi**

Tafadhali weka alama kwenye (√) chaguo lifaalo

1. Umri katika miaka katika miaka____________
2. Umri wa ndoa katika miaka_________
3. Nambari ya simu_________________
4. Hali ya ndoa

   (a)Ndoa ()

   (b) Wanaoishi pamoja ( )

5. Hali ya elimu

   (a) Hakuna ( )

   (b) Kiwango cha msingi hakijakamilika ( )

   (c) Kiwango cha msingi kimekamilika ( )

   (d) Sekondari au zaidi ( )

6. Hali ya ajira

   (a ) Walioajiriwa ( )

   (b) Wasio na kazi ( )

7. Dini

   (a)Mkristo ( )

   (b)Waislamu ( )

   (c) Wengine wanabainisha...

8. Kikundi cha kikabila

(a) Nyaturu ( )

(b) Kisukuma ( )

(c) Wengine hubainisha

9. Hali ya kiuchumi

(a) Tumia chini ya dola moja kwa siku ( )

(b) Tumia zaidi ya dola moja kwa siku ( )

10. Je, unamiliki redio?

(a) Ndiyo ( )

(b) Hapana ( )

11. Je, unamiliki simu ya mkononi?

(a) Ndiyo ( )

(b) Hapana ( )

12. Tabia ya kituo cha afya kilicho karibu

(a) Zahanati ( )

(b) Kituo cha afya ( )

(c) Hospitali ( )

13. Je, ni umbali gani wa kutembea hadi kituo cha afya kilicho karibu?

(a) Chini ya kilomita moja ( )

(b) Kilomita moja-kilomita 5 ( )

(c) Zaidi ya kilomita tano ()

14. Je, unahudumiwa na Bima ya afya (NHIF au CHF)?

(a) Ndiyo ( )

(b) Hapana ( )

15. Je, umewahi kusikia neno maandalizi ya kuzaliwa?

(a) Ndiyo ( ) (b) Hapana ( )

16. Ulisikia wapi kuhusu maandalizi ya kuzaliwa?

(a) Kutoka kwa mfanyakazi wa afya ( )

(b) Kutoka kwa vyombo vya habari ( )

(c) Kutoka kwa mwanafamilia ( )

(d) Wengine, taja…………

Sehemu B; Nia ya tabia ya wenzi wa kiume kuelekea kuhusika kwao katika ziara za utunzaji katika ujauzito na wanawake wao wajawazito

Maswali mengi katika utafiti huu yanatumia mizani ya kukadiria yenye nafasi 5; unatakiwa kuweka alama ya (√) kwenye kisanduku kinachoeleza vyema maoni yako ambapo SA= inakubali kabisa, A= inakubali, N= Si upande wowote, D= haikubaliani na SD = haikubaliani kabisa.

| SEHEMU | SA(5) | A(4) | N(3) | D(2) | SD(1) |
|---|---|---|---|---|---|
| B: Kuhusika kwa wanaume | | | | | |
| ***Mtazamo wa wenzi wa kiume kuelekea ushiriki wa wanaume;*** | | | | | |
| 1.Ikiwa mwenzi wa kiume atashiriki katika kuweka sehemu ya mkunga mwenye ujuzi, anafanya jambo jema na la manufaa. | | | | | |

| |
|---|
| 2.Ikiwa mwenzi wa kiume ataandamana na mwenzi wake wa kike mjamzito wakati wa kliniki ya ujauzito, anafanya jambo jema na la manufaa. |
| 3.Ikiwa mwenzi wa kiume atapima VVU na mwenzi wake wa kike wakati wa ujauzito, anafanya jambo jema na la manufaa. |
| 4.Iwapo mwanamume atafuatana na mwenzake wa kike wakati wa kujifungua, anafanya jambo jema lenye manufaa. |
| 5.Ikiwa mwenzi wa kiume ataandamana na mwenzi wake wa kike kwa uchunguzi baada ya kuzaa, anafanya jambo zuri na la manufaa |
| 6.Ikiwa mwenzi wa kiume atapima kaswende na mwenzi wake wa kike mjamzito wakati wa kliniki za wajawazito anafanya jambo jema ambalo ni la manufaa. |
| ***Tathmini ya kanuni za kibinafsi;*** |
| 7.Jamii inadhani mwenzi wa kiume lazima ashiriki katika kutenga fedha na vifaa vya kutumika katika dharura au wakati wa kujifungua. |

| |
|---|
| 8. Jamii inadhani mwenzi wa kiume lazima ashiriki katika maandalizi ya usafiri utakaotumika katika dharura au wakati wa kujifungua<br><br>9. Jamii inadhani mwenzi wa kiume anafaa kushiriki katika kumtambua mkunga mwenye ujuzi |
| 10.Jamii inafikiri kwamba mwenzi wa kiume anapaswa kuandamana na mwenzi wake wa kike mjamzito wakati wa kliniki za wajawazito |
| 11.Jamii inadhani mwenzi wa kiume anatakiwa kupima VVU na mpenzi wake wa kike mjamzito wakati wa ziara za ujauzito |
| 12.Jamii inafikiri kwamba mwenzi wa kiume lazima achunguze kaswende na mwenzi wake wa kike mjamzito wakati wa kutembelea katika ujauzito |
| 13.Jamii inafikiri kwamba mwanamume analazimika kuandamana na mwanamke mjamzito wakati wa kujifungua |
| 14.Jamii inafikiri kwamba mwenzi wa kiume anapaswa kuandamana |
| na mwenzi wake wa kike wakati wa uchunguzi wa baada ya kuzaa |
| ***Udhibiti wa Kitabia Unaojulikana;*** |
| 15.Kwa wewe kushiriki katika kuweka kando fedha na vifaa vya kutumika katika kesi ya dharura au wakati wa kujifungua ni rahisi na unaweza kufanya hivyo. |

| |
|---|
| 16.Kwa wewe kushiriki katika maandalizi ya usafiri wa kutumika katika kesi ya dharura au wakati wa kujifungua ni rahisi na unaweza kufanya hivyo. |
| 17.Kwa wewe kushiriki katika utambuzi wa mkunga mwenye ujuzi ni rahisi na unaweza kufanya hivyo |
| 18.Kwa wewe kuandamana na mwenzi wa kike mjamzito wakati wa kliniki za wajawazito ni rahisi na unaweza kufanya hivyo |
| 19.Kwa wewe kupima VVU/UKIMWI na mwenzi wako wa kike mjamzito wakati wa ziara za ujauzito haina shida na unaweza kufanya hivyo.<br><br>20.Kwa wewe kupima kaswende na mpenzi wako wa kike wakati wa kliniki za ujauzito ni rahisi na unaweza kufanya hivyo |
| 21.Kwa wewe kuandamana na mpenzi wako wa kike mjamzito wakati wa leba na kuzaa ni rahisi na unaweza kufanya hivyo. |
| 22.Kwa wewe kuandamana na mwenzi wako wa kike wakati wa ukaguzi baada ya kuzaa haina shida na unaweza kufanya hivyo |
